# Leprosy campaigns in Brazil: analysis of “Janeiro Roxo” impact using Google Trends

**DOI:** 10.1101/2023.10.04.23296467

**Authors:** Paulo Roberto Vasconcellos-Silva, Artur Custódio Moreira de Sousa

**Affiliations:** Oswaldo Cruz Institute/Fundação Oswaldo Cruz, Rio de Janeiro, RJ, Brazil; School of Medicine and Surgery, Federal University of the State of Rio de Janeiro/UNIRIO, Rio de Janeiro, RJ, Brazil; National School of Public Health, Oswaldo Cruz Foundation, Rio de Janeiro, RJ, Brazil

**Author notes:** These authors contributed equally to this work. Corresponding author (PRVS).

**Keywords:** Health communication, Internet and health, leprosy, health campaigns, Google Trends, leprosy early identification

## Abstract

“Janeiro Roxo” (Purple January) is a Brazilian campaign to raise awareness about leprosy and its early symptoms. The negative influence of the COVID-19 pandemic on institutional campaigns was especially relevant for the control of leprosy, if considered its status of neglected disease by the media. Google Trends (GT) is a free tool that tracks accesses to given terms on Google. Using GT we studied (i) impact of “Janeiro Roxo” on public interest on leprosy in the last 5 years; (ii) public interest during and after COVID-19 pandemic; (iii) patterns of public interest in self-identification of leprosy signs.

**Methods:** GT tracking follows the popularity of topics in time range in “Relative Search Volume” (RSV) – proxies of public interest in a particular subject on a scale from 0 to 100. We selected “HANSENÍASE” (HAN) and “HANSENÍASE SINTOMAS” (leprosy symptoms) (H.SIN) to estimate public interest in leprosy and “self-diagnosis” during 261 weeks (August 2018 to 2023). Polynomial trend lines estimate trends over the period. Weekly RSV, monthly and annual means were compared using Analysis of Variance (ANOVA).

**Results:** In 261 weeks: higher RSV to “HANSENÍASE” (HAN) compared to “HANSENÍASE SINTOMAS” (H.SIN); highest Monthly Means (MM) in January months (2019 to 2023) for both HAN and H.SIN.

*Pandemic effect in “Janeiro Roxo”:* decrease in January MM 2021 compared to 2020 (24% to HAN, 25% to H.SIN). Both HAN/H.SIN reached pre-pandemic levels in January 2022/2023.

*Breakpoints:* (points of abrupt change which influences trend lines) in the 26th week (February 2019); in the 55^th^ and 213^th^ week (September 2019 and 2022). Trend line (HAN): upward curve between 33^rd^-45th week (April to June 2019); pandemic downward trend between 120^th^-136th week (December 2020 to March 2021); upward trend curve between 220^th^-240th week (November 2022 to March 2023).

**Conclusion:** “Janeiro Roxo” and in-person activities, among other media stimuli, have a relevant impact in terms of public interest in leprosy, even after their interruption due to pandemic. Considering their impacts on national searches, regional campaigns should be considered very successful. RSV to HAN may be due to “reactive searches” motivated by transient stimuli from campaigns, in an ephemeral and impersonal way. “Proactive searches” to H.SIN may be associated to those interested - in a personal way - in early symptoms and self-diagnosis. Internet-based campaign evaluation programs could be useful in integrating vulnerable regions and exposing their needs for assistance services.

## Introduction

Leprosy is a neglected disease that remains endemic as a major public health problem in large areas of the planet. India, Brazil and Indonesia are the countries that reported the most new cases, corresponding to 74.5% of the global total [1]. Although it is still a neglected disease, prevalence has decreased substantially, from more than 5 million cases in the 1980s to 133,802 cases in 2021 [1]. This may be due to institutional efforts, with investments in campaigns for early identification and treatment. “Janeiro Roxo” (Purple January) is a Brazilian campaign coordinated by Ministry of Health, State Health Departments and Municipalities to raise awareness about leprosy and its early symptoms [2]. In “Janeiro Roxo”, health authorities and non-governmental organizations (NGOs) emphasize the stigma associated with leprosy to reintegrate those affected and disseminate information about the disease and its treatment. During the campaign, several events are held across the country: lectures, seminars, workshops and educational activities in schools and communities. Insertions on media and social networks are also frequent.

The increased media focus on actions aimed at controlling the COVID-19 pandemic may have potentially reallocated resources and public attention away from health awareness campaigns concerning other diseases [3]. In recent years, several articles have described the negative influence of the pandemic [4–6] on institutional campaigns planned to inform society about early identification of diseases [7-8]. This aspect is especially important for the control of leprosy, mostly when considering its status as a neglected disease by the media [9-10]. Organizers of “Janeiro Roxo” were apprehensive about the potential negative impact of pandemic on future campaigns and new leprosy cases. Public health planners feared a setback, foreseeing a rise in new cases due to insufficient interest in leprosy early symptoms.

Several works have focused on improving the design and evaluation of institutional campaigns similar to “Janeiro Roxo”. Studies have proposed theoretical frameworks for effective health campaigns design [11-13], examined approaches to evaluating outcomes [14], user acceptance and new suggestions of media– based interventions [15-17]. Currently, the problem persists without plainly efficient tools to evaluate results and chart new directions [18]. In this context, Google Trends (GT) methodology has been used as a public interest proxy in health-related phenomena, identifying trends and patterns of accesses to various conditions in different circumstances [19-25].

To advance understanding of the potential for Google Trends to gauge online interest in leprosy control and identify negative drivers of changes in search volume, this study uses a time series analysis on GT (2018 to 2023) to investigate: (i) trends of public interest in terms associated to leprosy during “Janeiro Roxo” campaigns; (ii) patterns on this public interest during COVID-19 pandemic; (iii) patterns of interest in self-identification of leprosy signs. We tracked Google searches targeting “HANSENÍASE” (leprosy) as a general term. Considering that the main objective of the campaign is to raise awareness about leprosy early identification, we also analyzed a specific subset of searches targeting symptoms - “HANSENÍASE SINTOMAS” (leprosy symptoms).

## Methodology

Google Trends (GT) is a free tracking system based on algorithms that measure access to given terms on Google. The open tool allows the user to customize and obtain volumes of search terms to infer trends of public interest in a given topic at a given time. GT tracking estimates the relative popularity of topics in time range, but does not provide an absolute count of searches; rather, GT normalize data for the overall number of searches on a scale from 0 (search volume <1% of the peak volume) to 100 (peak of public interest). Results are presented as weekly “Relative Search Volume” which are, by definition, always less than 100 - a proportion compared with the highest search volume (RSV:100) [26]. RSV oscillations and trends are interpreted as proxy of public interest in a particular search issue, at a certain time point. According to literature, this approach corrects results for population size and Internet access [25].

To represent collective interest in leprosy, the following keywords were compared: “HANSENÍASE”; “HANSENIASE” (with and without an acute accent); “LEPRA”; “LEPROSE”; “DOENÇA DE HANSEN”; “MAL DE HANSEN”; and “MORFEIA”. We selected “HANSENÍASE” (with acute accent) due to the highest search volume in the period, compared to the others. The term “HANSENÍASE SINTOMAS” (leprosy symptoms) was tracked to estimate lay public interest concerning “self-diagnosis”, as referred by literature [27-28]. We used “Brazil” as country and category “Health” to produce a 5 years’ time series (08/26/2018 to 08/20/2023) with 261 weeks. We downloaded data in CSV format to plot a graph with 261 RSVs for both keywords. Polynomial trend lines (sixth order) were added up to estimate trends over the period. Weekly RSV, monthly and annual means were compared using Analysis of Variance (ANOVA) to analyze differences among means.

### Ethical approval

The current research analyzed retrospectively open and anonymized data. According to Resolution 510/2016 of the Brazilian National Health Council, all research involving public domain data and unidentified participants does not require ethical assessment by the Research Ethics National Council system (CONEP). The authors assert that all procedures contributing to this work comply with the ethical standards of the relevant national and institutional committees on human experimentation and with the Helsinki Declaration of 1975, as revised in 2008.

## Results

During the 261 weeks, Google Trends registered a significantly higher volume of hits to keyword “HANSENÍASE” (HAN) and greater fluctuations if compared to “HANSENÍASE SINTOMAS” (H.SIN). Concerning the evolution of HAN and H.SIN Annual Means from 2018-2023 (without January in 2018): increase in RSVs to HAN/H.SIN from 2018-2019, followed by reductions in 2020-2021 (24% to HAN and 32.5% to H.SIN), returning to pre-pandemic levels in 2022 and 2023 (Table 1, Fig 1 and 2).

**Table 1.**
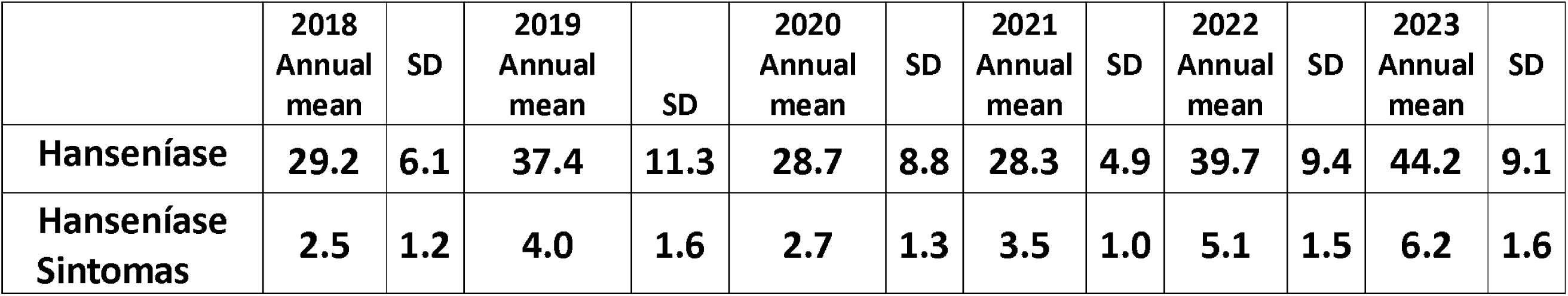
Annual means of “HANSENÍASE” and “HANSENÍASE SINTOMAS” (2018 to 2023).

|  | 2018<br>Annual<br>mean | SD | 2019<br>Annual<br>mean | SD | 2020<br>Annual<br>mean | SD | 2021<br>Annual<br>mean | SD | 2022<br>Annual<br>mean | SD | 2023<br>Annual<br>mean | SD |
| --- | --- | --- | --- | --- | --- | --- | --- | --- | --- | --- | --- | --- |
| Hanseníase | 29.2 | 6.1 | 37.4 | 11.3 | 28.7 | 8.8 | 28.3 | 4.9 | 39.7 | 9.4 | 44.2 | 9.1 |
| Hanseníase Sintomas | 2.5 | 1.2 | 4.0 | 1.6 | 2.7 | 1.3 | 3.5 | 1.0 | 5.1 | 1.5 | 6.2 | 1.6 |

**Figure 1.**
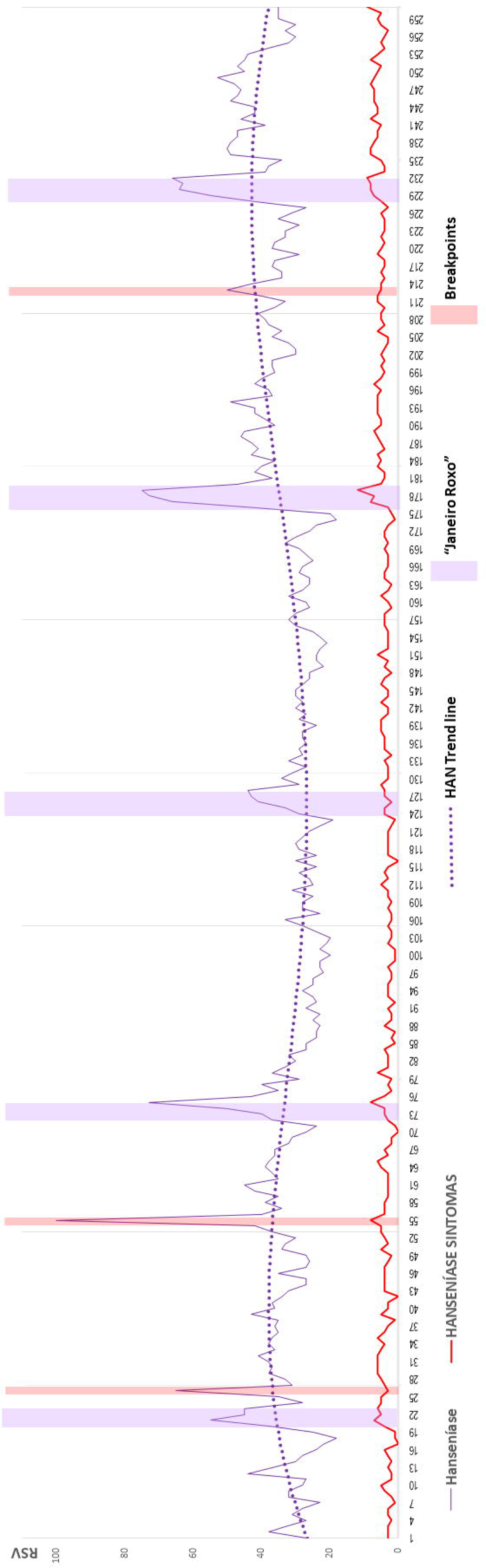
Annual means of “HANSENÍASE” and “HANSENÍASE SINTOMAS” (2018 to 2023).

**Figure 2.**
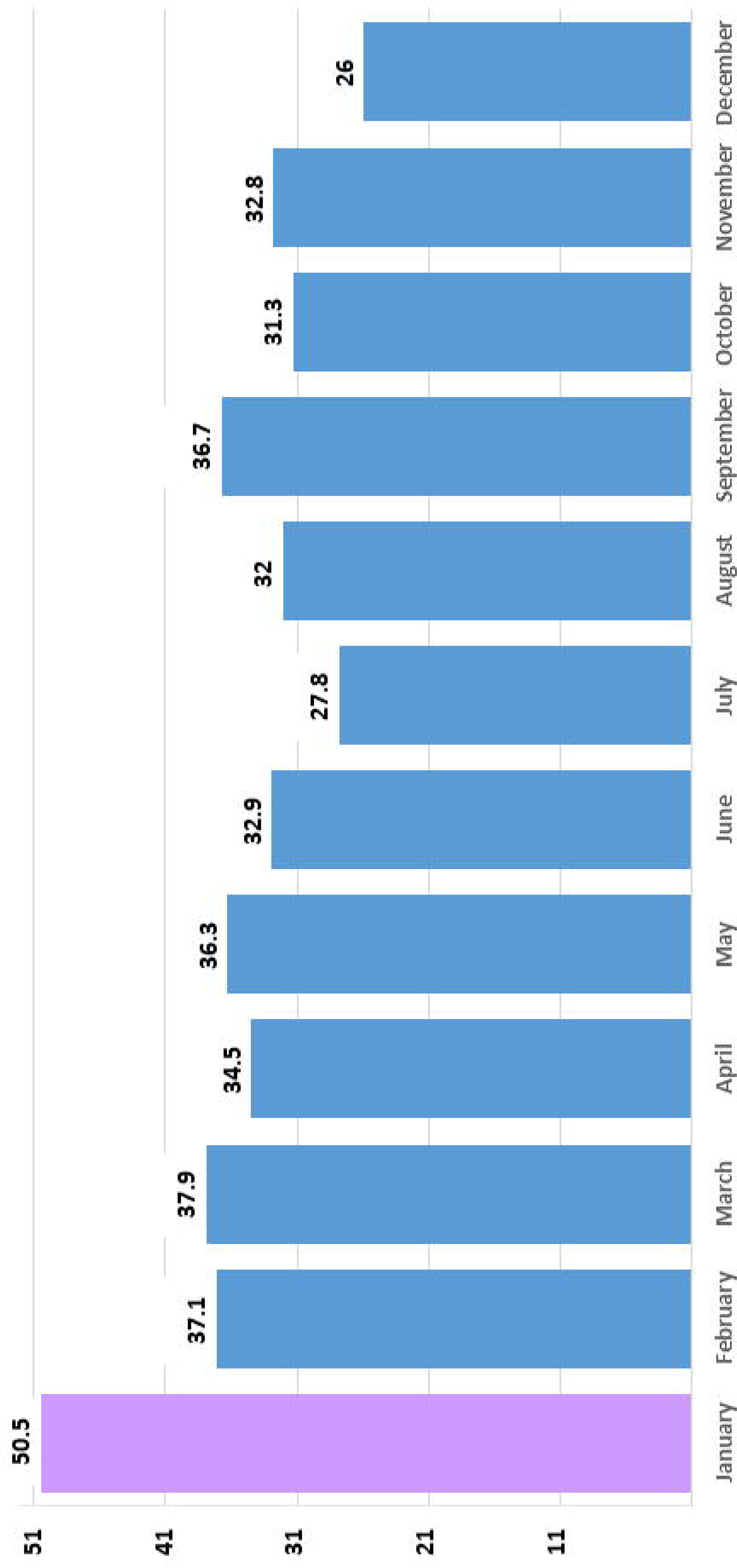
Evolution of the 261 weekly RSV to “HANSENÍASE” and “HANSENÍASE SINTOMAS” (2018-2023); “Janeiro Roxo”, breakpoints and HAN trend line.

### Monthly means in “Janeiro Roxo” and pandemic

The highest Monthly Means (MM) were observed in January months from 2019 to 2023, for both HAN and H.SIN (Table 2 and Fig 3 and 4).

**Table 2.**
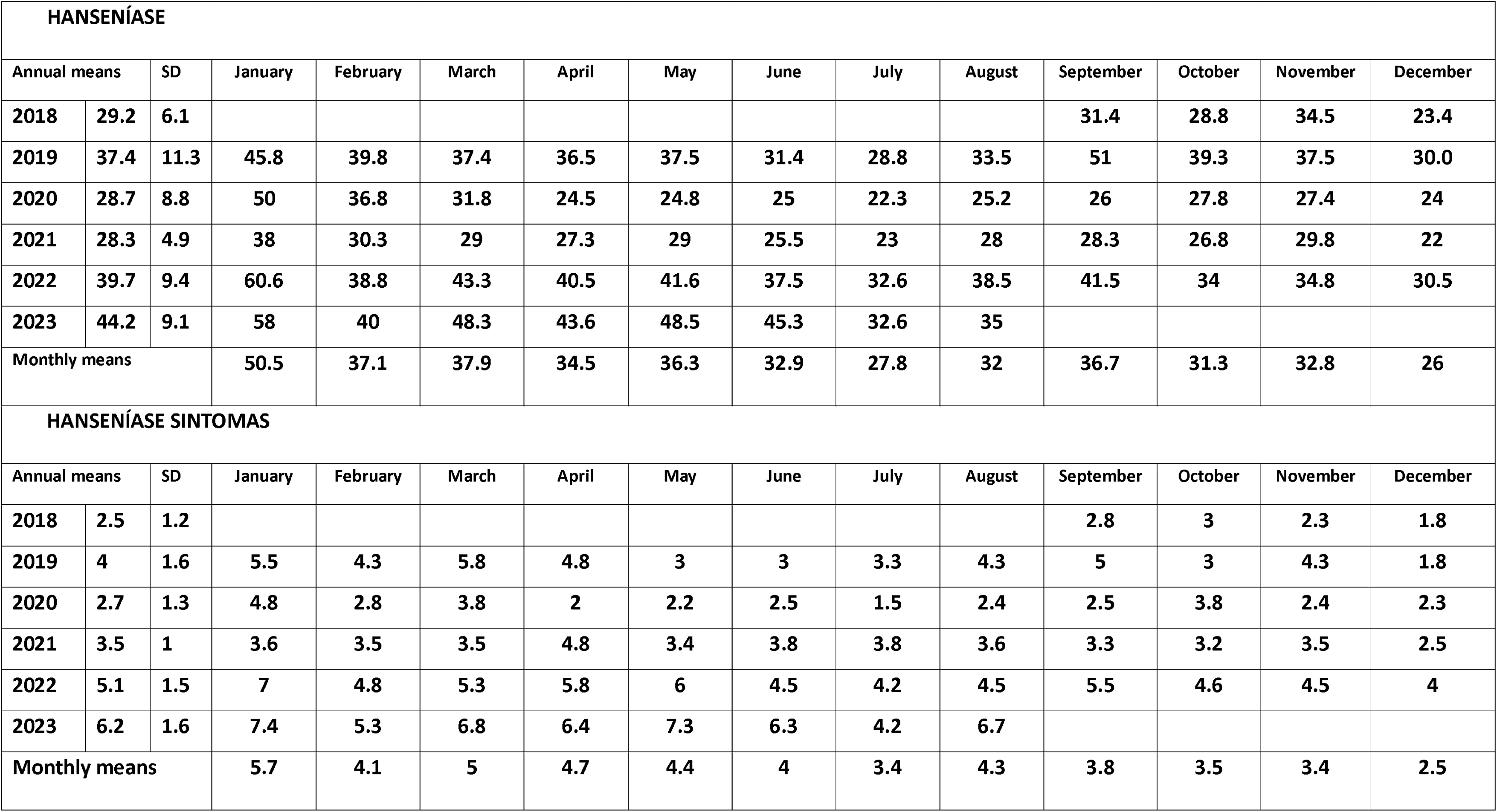
Monthly means and Annual means (2018 to 2023) to “HANSENÍASE” e “HANSENÍASE SINTOMAS”.

**Figure 3.**
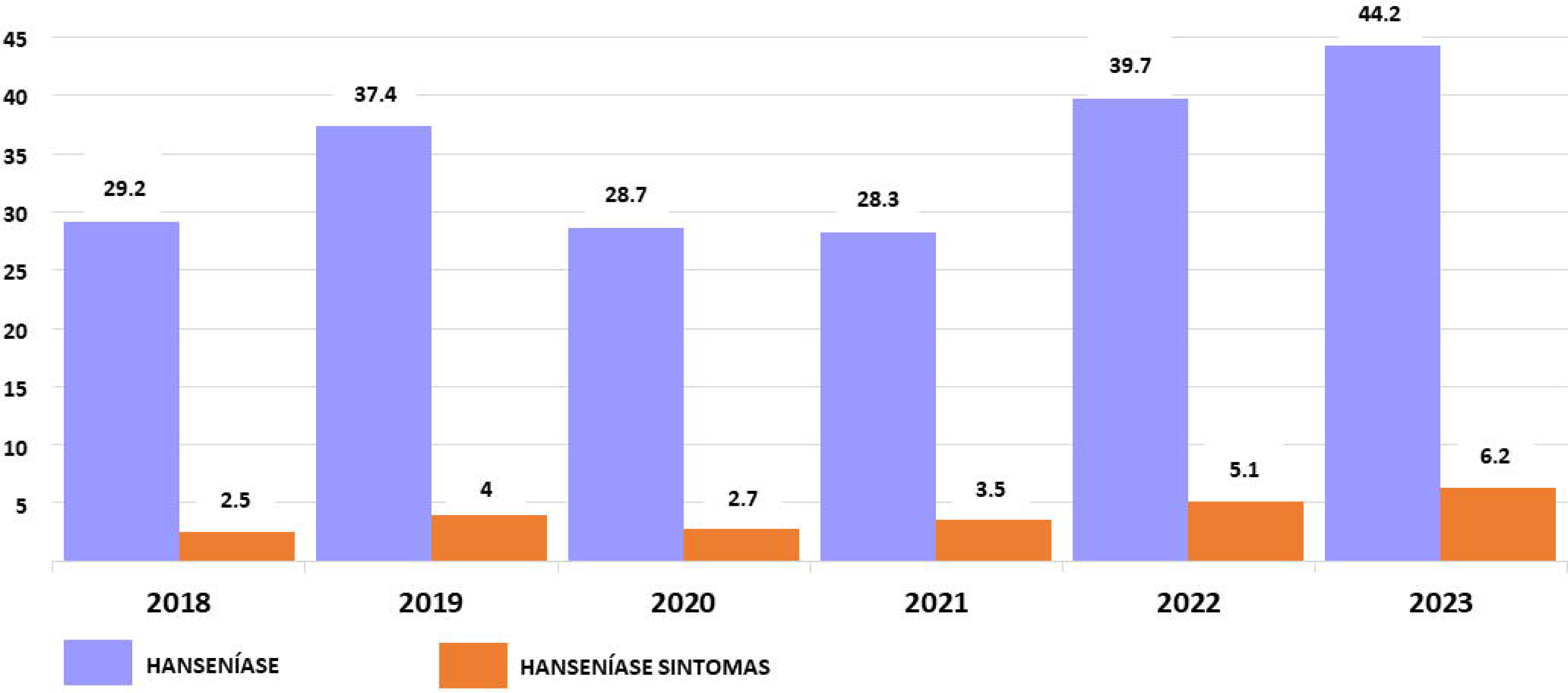
“Janeiro Roxo” Monthly means (2019 to 2023) compared to other months - “HANSENÍASE”.

**Figure 4.**
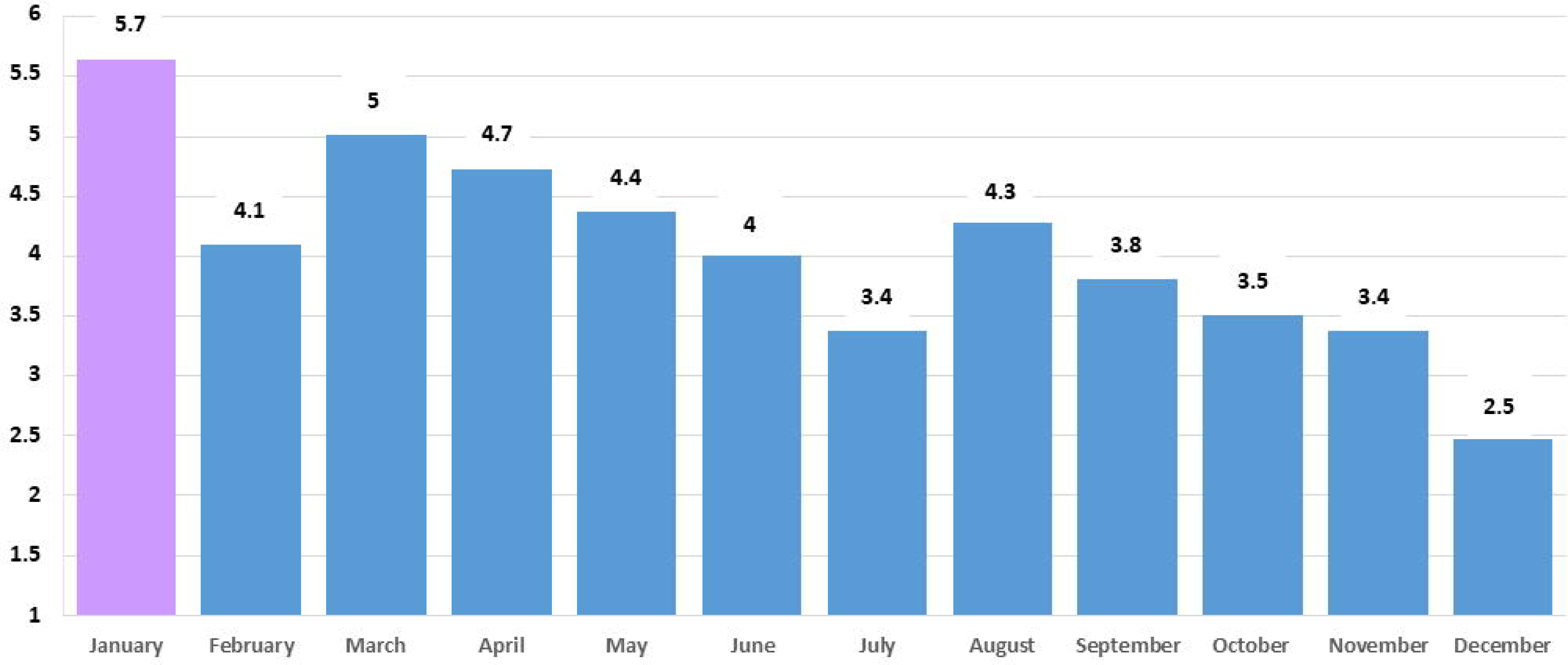
“Janeiro Roxo” Monthly means (2019 to 2023) compared to other months - “HANSENÍASE SINTOMAS”.

Concerning the pandemic effect in “Janeiro Roxo”: to HAN there was a 24% decrease in January MM 2020 (before pandemic) compared to January MM 2021; to H.SIN there was a 25% decrease in the same period (Table 2). Both HAN/H.SIN reached pre-pandemic levels in January 2022/2023, as shown in Table 2.

### RSV Breakpoints

A first breakpoint (i.e., points of abrupt change which influences trend lines) were observed in the 26th week only to HAN - RSV:65 (from 17 to 24/02/2019) – 2019 AM: 37.4 (SD:11.3) (red area in Fig 2). The highest peak in searches to HAN was observed in the 55^th^ week (from 08 to 15/09/2019) with RSV:100 - 2019 AM:37.4 (SD: 11.3) and H.SIN - RSV:8 - 2019 AM:4 (SD:1.6). In the 213th week (from 18 to 25/09/2022) was observed a new breakpoint only to HAN (RSV: 50 – 2022 AM: 39.7, SD: 9.4) (red area in Fig 2).

### Trend lines

RSV trend lines to HAN show an upward curve to the 1st plateau between the 33rd and 45th week (April to June 2019), related to higher MM from 2018 to 2019. There was a downward trend between the 120th and 136th week (December 2020 to March 2021) reflecting the reduced pandemic RSVs; and a new upward trend is present between the 220th and 240th week (November 2022 and March 2023) (Fig 2). RSV trend lines for H.SIN are not evident due to lower RSV volume and small fluctuations.

## Discussion

At the national level, we found a relevant disproportion of accesses to HAN compared to H.SIN, although we must consider that RSVs to H.SIN are a subset of the searches to HAN. It is important to note that, proportionally, the pandemic had a more negative influence on searches for symptoms (32.5% decrease to H.SIN) compared to searches for leprosy (24% decrease to HAN). These differences may be due to factors associated to the nature of searches. Researchers, technical public and lay public, in general, may be interested (theoretically or just by simple curiosity) in the disease itself, its origins, contamination vectors, treatments, among other aspects. This behavior is defined by literature as “reactive searches” [29-30], motivated by transient stimuli from campaigns and news in the media, in an ephemeral and impersonal way. On the other hand, “proactive searches” may be performed by those who have already identified some signs of the disease in their body. They may be interested - in a personal way - in early symptoms when using “Doctor Google” to self-diagnosis [27-28]. Important to stress that leprosy campaigns are primarily aimed to those worried about symptoms, perhaps indicating the need for immediate technical assessment. Usually “Doctor Google” is a first step to guide the next one. Thus, people interested in their own health conditions would carry out a “proactive search” motivated by campaigns or any other type of frequent stimulus in the media. [29].

### “Janeiro roxo” campaigns

Another relevant finding refers to the reactions triggered by “Janeiro Roxo” campaigns. Significant increases in searches were observed in January, for both HAN and H.SIN - even during the pandemic period, in a lower proportion (Table 2). The “pandemic effect” is suggested by the trend lines to HAN from the 80th (March 2020) to the 175th week (end of 2021) (Fig 2). According to RSV oscillations to HAN during “Janeiro Roxo”, an increase can be observed in 2019 - 2020 (January MM: 45.8 – 50); until the “COVID-19 eclipse”, period of the out-of-control pandemic (2021 January MM:38); with recovery in 2022 / 2023 (January MM: 60.6 / 58) surpassing Annual Means - 2022 AM: 39.7 (SD 4.9) / 2023 AM: 44.2 (SD: 9.1). RSV to H.SIN, although on a lower scale, maintained the same pattern (Table 2, Fig 2).

### RSV Breakpoints

Regarding breakpoints, we adjusted GT search terms to “Brazil”. However, it is relevant to consider Brazilian continental dimensions and their regional / epidemiological peculiarities. GT parameters adjusted only for the Brazilian North and Northeast regions (where the highest numbers of cases are reported) would be sensible to regional impacts of local campaigns – in particular those that take place after January. Since September 2016, the “State Day for Awareness, Prevention and Combat of Leprosy” has been organized as a regional campaign in Piauí (Brazilian Northeastern state) [31]. Health authorities and NGOs organize intense programming to influence the inhabitants of Piauí, students and health managers. These efforts should be considered very successful, considering their impacts on national searches - in the 55th week, between 09/08/2019 and 09/15/2019, RSVs to HAN reached its peak RSV:100 (contrast to 2019 AM:37.4 - SD: 11.3) and H.SIN RSV:8 (2019 AM:4 - SD: 1.6) (Table 2). During the period studied, regional campaigns in September (2018 – 2023) achieved high RSVs, although not reaching the same level as “Janeiro Roxo” - a national traditional event. In 2020/2021 “State Day for Awareness” campaigns did not take place, which is reflected in the lower RSVs compared to 2020/2021 AM (Table 2). Therefore, consistent with GT logic, the 2019 “State Day for Awareness” campaign should be considered a gold standard on which ensuing campaigns should be based.

Perhaps influenced by previous “Janeiro Roxo”, in February 2019 several academic publications and newspaper articles were disseminated, among other media stimuli [32-37]. The 26th week breakpoint registered RSV:65 to HAN (February 2019), significantly higher than MM:39.8 (SD: 17.1) and AM: 37.4 (SD:11.3). The same was not observed to H.SIN (RSV: 3) (Fig 2), perhaps because these publications did not emphasize information concerning symptoms, inherent motivation to searches to H.SIN.

## Conclusion

As usually happens in “Pink October” [38] and “Blue November” [39], institutional campaigns such as “Janeiro Roxo” seem to have a relevant impact that can be identified and quantified by GT algorithmics. Brazilian campaigns draw attention to the relevance of festivities and in-person activities that support these interventions, as described in the literature [40]. As shown above, after the interruption of face-to-face campaigns there were signs of a “resurgence” of social interest in leprosy and its early identification. Important to stress that understanding the relative popularity of topics, rather than attempting to assess absolute numbers (essential to epidemiological approaches) could assist the comprehension of communication approaches singularity. In other words, although the method does not determine unequivocal linear causal relationships, it can point out golden standards and useful evidence for building models that should be considered. Geosegmentation in search filters could be a useful alternative for large countries like Brazil. Perhaps it will allow researchers to identify the most vulnerable sub-regions, with a high incidence of new cases and frequent Google searches to first signs of leprosy. Additionally, the low costs involved, simple analysis methodology and immediately applicable results could help different regions of the world to develop alternatives for planning locally effective campaigns. Internet-based campaign evaluation programs could therefore be useful in integrating isolated and vulnerable regions and exposing their needs for assistance services.

## Limitations

Google Trend’s limitations must be taken into account in its interpretation, including limited validity in areas with low Internet penetration, lack of information on absolute search volumes, and limited transparency in its algorithm. Although Internet access is precarious in several regions of Brazil, it would be interesting to consider the data collected here as a valid proxy, capable of extrapolation to inhabitants of higher risk areas.

## Supporting information

Supplemental CSV file

## Data Availability

All relevant data are within the manuscript and its Supporting Information files.

## Acknowledgements

MORHAN/Movement for the Reintegration of People Affected by Leprosy (NGO) for their help with data collection.

## Supporting information

S1. Table 1 – Annual means of “HANSENÍASE” and “HANSENÍASE SINTOMAS” (2018 to 2023).

S2. Table 2 – Monthly means and Annual means (2018 to 2023) to “HANSENÍASE” e “HANSENÍASE SINTOMAS”.

S1. XLSX. Spreadsheet with base data.

S2. CSV. Spreadsheet downloaded from Google Trends.

