## Supplemental TABLE 1 file for "Leprosy campaigns in Brazil: analysis of “Janeiro Roxo” impact using Google Trends"

|  | **2018  Annual mean** | **SD** | **2019**  **Annual mean** | **SD** | **2020  Annual mean** | **SD** | **2021  Annual mean** | **SD** | **2022  Annual mean** | **SD** | **2023  Annual mean** | **SD** |
| --- | --- | --- | --- | --- | --- | --- | --- | --- | --- | --- | --- | --- |
| **Hanseníase** | **29.2** | **6.1** | **37.4** | **11.3** | **28.7** | **8.8** | **28.3** | **4.9** | **39.7** | **9.4** | **44.2** | **9.1** |
| **Hanseníase Sintomas** | **2.5** | **1.2** | **4** | **1.6** | **2.7** | **1.3** | **3.5** | **1** | **5.1** | **1.5** | **6.2** | **1.6** |

**Table 1 – Annual means of “HANSENÍASE” and “HANSENÍASE SINTOMAS” (2018 to 2023).**
