## Supplemental TABLE 2 file for "Leprosy campaigns in Brazil: analysis of “Janeiro Roxo” impact using Google Trends"

| **HANSENÍASE** | | |  |  |  |  |  |  |  |  |  |  |  |  |
| --- | --- | --- | --- | --- | --- | --- | --- | --- | --- | --- | --- | --- | --- | --- |
| Annual means | | SD | **January** | **February** | **March** | **April** | **May** | **June** | **July** | **August** | **September** | **October** | **November** | **December** |
| **2018** | **29.2** | 6.1 |  |  |  |  |  |  |  |  | 31.4 | 28.8 | 34.5 | 23.4 |
| **2019** | **37.4** | 11.3 | 45.8 | 39.8 | 37.4 | 36.5 | 37.5 | 31.4 | 28.8 | 33.5 | 51 | 39.3 | 37.5 | 30 |
| **2020** | **28.7** | 8.8 | 50 | 36.8 | 31.8 | 24.5 | 24.8 | 25 | 22.3 | 25.2 | 26 | 27.8 | 27.4 | 24 |
| **2021** | **28.3** | 4.9 | 38 | 30.3 | 29 | 27.3 | 29 | 25.5 | 23 | 28 | 28.3 | 26.8 | 29.8 | 22 |
| **2022** | **39.7** | 9.4 | 60.6 | 38.8 | 43.3 | 40.5 | 41.6 | 37.5 | 32.6 | 38.5 | 41.5 | 34 | 34.8 | 30.5 |
| **2023** | **44.2** | 9.1 | 58 | 40 | 48.3 | 43.6 | 48.5 | 45.3 | 32.6 | 35 |  |  |  |  |
| **Monthly means** | | | **50.5** | **37.1** | **37.9** | **34.5** | **36.3** | **32.9** | **27.8** | **32** | **36.7** | **31.3** | **32.8** | **26** |
| **HANSENÍASE SINTOMAS** | | | |  |  |  |  |  |  |  |  |  |  |  |
| Annual means | | SD | **January** | **February** | **March** | **April** | **May** | **June** | **July** | **August** | **September** | **October** | **November** | **December** |
| **2018** | **2.5** | 1.2 |  |  |  |  |  |  |  |  | 2.8 | 3 | 2.3 | 1.8 |
| **2019** | **4** | 1.6 | 5.5 | 4.3 | 5.8 | 4.8 | 3 | 3 | 3.3 | 4.3 | 5 | 3 | 4.3 | 1.8 |
| **2020** | **2.7** | 1.3 | 4.8 | 2.8 | 3.8 | 2 | 2.2 | 2.5 | 1.5 | 2.4 | 2.5 | 3.8 | 2.4 | 2.3 |
| **2021** | **3.5** | 1 | 3.6 | 3.5 | 3.5 | 4.8 | 3.4 | 3.8 | 3.8 | 3.6 | 3.3 | 3.2 | 3.5 | 2.5 |
| **2022** | **5.1** | 1.5 | 7 | 4.8 | 5.3 | 5.8 | 6 | 4.5 | 4.2 | 4.5 | 5.5 | 4.6 | 4.5 | 4 |
| **2023** | **6.2** | 1.6 | 7.4 | 5.3 | 6.8 | 6.4 | 7.3 | 6.3 | 4.2 | 6.7 |  |  |  |  |
| **Monthly means** | | | **5.7** | **4.1** | **5** | **4.7** | **4.4** | **4** | **3.4** | **4.3** | **3.8** | **3.5** | **3.4** | **2.5** |

**Table 2 – Monthly means and Annual means (2018 to 2023) to “HANSENÍASE” e “HANSENÍASE SINTOMAS”.**
